# Social Inequities in Transportation Noise Exposures in the United States: Urbanicity Modifies that Relationship Between Social Vulnerability and Noise Exposure

**DOI:** 10.1101/2025.08.15.25333812

**Authors:** Ching-Hsuan Shirley Huang, Elena Austin, Edmund Seto

**Affiliations:** Department of Environmental and Occupational Health Sciences, School of Public Health, University of Washington, 98105

**Keywords:** social vulnerability index (SVI), environmental justice, transportation noise exposure, urbanization, principal component analysis (PCA), principal component regression (PCR)

## Abstract

Prior research has explored the unequal distribution of environmental hazards; however, little is known about the relationship between social vulnerability and transportation noise exposure in the U.S., particularly in relation to urbanization levels. We sought to examine the relationship between the Social Vulnerability Index (SVI) and transportation noise exposure across the U.S. at the census-tract level and assessed the moderating effects of urbanization. Multivariate linear regression models were used to assess the relationship between SVI – a composite index consisting of thematic indicators for socioeconomic status (SES), household characteristics, racial/ethnic minority status, and housing/transportation factors – and population-weighted transportation noise exposure at the census-tract level. An interaction term was used in the model to assess the moderating effects of urbanization. Principal component analysis (PCA) and regression (PCR) were employed to extract the features that correlate with specific SVI themes and examine their relationship to transportation noise exposure. Findings from PCA and PCR reveal that in the urban area context, census tracts that score high on all four vulnerability themes of the SVI are positively associated with transportation noise exposure, while an opposite relationship is observed for census tracts that score high in vulnerability with respect to racial/ethnic minority status and household characteristics, but not particularly high in vulnerability for SES or housing/transportation factors, regardless of the urbanization level. Our study underscores the impact of distinct social vulnerability components on transportation noise exposure, revealing variations across different urbanization strata.

**Highlights:**

- Social vulnerability and urbanization levels influence transportation noise exposure.
- Urban areas show positive associations between SVI and noise; rural areas show inverse trends.
- PCA and PCR reveal distinct SVI components linked to transportation noise exposure patterns.
- High SVI urban tracts experience elevated noise; rural racial/ethnic minority tracts are less exposed.
- Findings inform targeted policies addressing environmental justice and noise mitigation strategies.

## 1. Introduction

Transportation noise has been increasingly considered as a significant public health concern. The European Environment Agency (EAA) estimated that in 2018, over 100 million people in Europe were exposed to day-night-evening noise levels (Lden) over 55 dB, leading to high annoyance and sleep disturbances, particularly due to aircraft, rail, and roadway noise (World Health Organization, 2018). In the U.S., nationwide studies have reported that an estimated of 94 million individuals were exposed to transportation noise levels surpassing 45 dB in 2020 (Seto and Huang, 2023b), with an estimated 7.8 million (2.40% of the total population), 5.2 million (1.60% of the total population), and 7.9 million (2.43% of the total population) individuals highly annoyed by aviation, rail, and roadway noise, respectively (Huang and Seto, 2023). In addition to high annoyance and sleep disturbance, exposure to noise has been found to be associated with increased risk of cardiovascular diseases (Van Kempen et al., 2018), anxiety and depression (Beutel et al., 2016), impacts on children’s learning (Stansfeld et al., 2005), and low birth weight children (Argys et al., 2020).

Urbanization, marked by swift development in transportation infrastructure and population density, brings forth not only progress but also challenges. Between 2010 and 2020, the population living in urban areas of the U.S., increased by 6.4%, now accounting for 80% of the nation’s population (U.S. Census Bureau., 2022). This trend is associated not only with economic and social advancement but also with escalating levels of environmental pollution, including air and noise pollution, impacting various dimensions of human well-being. Research has also indicated a pronounced unequal distribution of these environmental pollution and its associated burden (Wang, 2018, Liu et al., 2021, Young et al., 2012, Woo et al., 2019, Casey et al., 2017, Collins et al., 2020). Notably, fewer investigations have delved into the socio-economic disparities of community noise exposure at a nationwide scale in the U.S (Casey et al., 2017, Collins et al., 2020). Recent research has revealed its disproportionate impact on minority populations, with Asians highly annoyed by roadway noise, and Black and Hispanic populations most affected by rail and aviation noise, respectively (Huang and Seto, 2023). Socioeconomic status also plays a pivotal role in shaping the impact of transportation noise exposure on communities. Areas characterized by lower socioeconomic conditions, often indicative of poverty, limited access to educational opportunities, and insufficient healthcare infrastructure, tend to exhibit heightened susceptibility to the adverse impacts of environmental hazards. These characteristics, often combined through the concept of social vulnerability, has been widely studied and various measures based on different sociodemographic and socioeconomic factors have been established (Cutter et al., 2003, Flanagan et al., 2011, Felsenstein and Lichter, 2014, Aroca-Jiménez et al., 2020, Lee, 2014).

Most U.S. based noise studies have only considered equity indicators (e.g., racial/ethnic minority status or SES indicators) as independent risk factors (Casey et al., 2017, Collins et al., 2019) rather than from the concept of holistic social vulnerability. In the U.S., the Centers for Disease Control, Agency for Toxic Substances and Disease Registry (CDC/ATSDR) developed the Social Vulnerability Index (SVI), providing a systematic framework for quantifying a community’s capacity for disaster preparedness and response, and exposures to hazards and community-level stressors (Centers for Disease Control and Prevention/ Agency for Toxic Substances and Disease Registry/ Geospatial Research, 2020). The CDC/ATSDR SVI integrates 16 sociodemographic factors/attributes across four themes, including socioeconomic status, household characteristics, race and ethnic minority status, and housing type and transportation, providing a holistic assessment of social vulnerability by considering various dimensions that collectively contribute to the overall vulnerability of communities. The summation of the factors explicitly acknowledges that these factors act cumulatively to characterize the social vulnerability of communities. The selection of these themes were based on extensive literature review (Flanagan et al., 2011). The SVI was formulated by ranking 16 sociodemographic factors/attributes for each census tract from zero to one and summing the rankings, and followed by a re-ranking procedure from the most to least vulnerable. Due to its ease of use and comprehensive nature (Tarling, 2017), the CDC/ATSDR SVI has been widely applied in different research studies to investigate the role of social vulnerability and the burden of natural disasters and environmental hazards.

In this study, our primary objectives are to explore the relationship between social vulnerability and transportation noise exposure in the U.S., while assessing the role of urbanization level in moderating this relationship. Furthermore, we aimed to explore the underlying sociodemographic factors contributing to the CDC/ATSDR SVI and examine their connection to transportation noise exposure. Leveraging a national transportation noise exposure dataset from a prior study (Seto and Huang, 2023b) and the CDC/ATSDR SVI, we conducted a spatial analysis to map the distribution of population-weighted transportation noise exposure and CDC/ATSDR SVI across the U.S. Our approach involved employing multivariate and principal component regression to model and understand the relationships between these variables.

## 2. Materials and methods

### 2.1 Census-tract level transportation noise exposure data

We obtained the census-tract level national transportation noise exposure (NTNE) data from a prior study (Seto and Huang, 2023b). To summarize, the dataset was generated by overlaying the most recent (2020) U.S. Bureau of Transportation Statistics’ (BTS) National Transportation Noise Map (NTNM) spatial raster files of noise levels associated with aviation, rail, and roadway traffic noise at 30 m spatial resolution with the spatial population estimates from the 5-year (2016 - 2020) American Community Survey (ACS) at the census block-group level. The exposed population of each census block group was overlaid with LAeq noise categories of 45 - 50, 50 - 60, 60 - 70, 70 - 80, 80 - 90, and 90+ dBA LAeq (greater than or equal to the lower bound and less than the upper bound applied to each category) and aggregated to the census tract level.

The BTS NTNM files were organized by state and incorporated as pooled or source-specific (i.e., aviation, rail, or roadway source) data. Detailed information regarding the methodologies employed for noise level modeling can be found in the BTS documentation (U.S. Bureau of Transporation Statistics, 2020). In the current analysis, we retrieved the NTNE data combining all sources of transportation, including aviation, rail, and roadway. For each census tract, we computed the population-weighted noise exposure using the midpoint level of each LAeq noise categories:

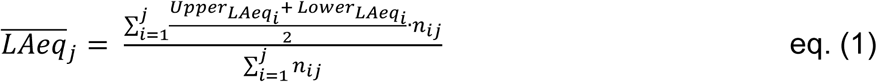

where 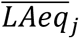 is the population-weighted noise exposure level (LAeq) of census tract *j*; *Upper_LAeq_i__* and *Lower_LAeq_i__* are the upper bound and lower bound of LAeq noise category *i*, respectively (LAeq); and *n_ij_* is the exposed population numbers in noise category *i* of census tract *j*. For LAeq noise category 90+, 90 dBA LAeq was input as the midpoint level for calculation.

To evaluate the role of urbanization in population noise exposure, we aggregated the data by Urbanized Area (referred to as “UA” hereafter) and the top 5% most populous Urbanized Area (referred to as “HUA” (Highly Urbanized Area) hereafter). The Urban Area classification is defined by the U.S. Census Bureau as territories with 50,000 or more residents, representing densely developed areas that include residential, commercial, and other non-residential urban land uses. The HUA in our analysis included a total of 24 Urban Areas, each with over 2,000,000 residents.

### 2.2 Census-tract level social vulnerability data

The census-tract level CDC/ATSDR SVI data for the year 2020 were obtained from the CDC website (Centers for Disease Control and Prevention/ Agency for Toxic Substances and Disease Registry/ Geospatial Research, 2020). The methodology for developing the CDC/ATSDR SVI was described by Flanagan et al. (Flanagan et al., 2011). The CDC/ATSDR SVI incorporates 16 factors across four themes, including socioeconomic status, household characteristics, racial and ethnic minority status, and housing type and transportation. These variables (**Table 1**) were sourced from the American Community Survey (ACS). For each census tract, the SVI provides both raw scores and percentile rankings. The raw scores are calculated by summing the percentile values of variables within each theme, and then across all four themes, to generate the overall SVI score. While the raw scores can exceed 1 due to this summation process, the percentile rankings are subsequently computed to standardize the overall SVI scores on a scale from 0 (lowest vulnerability) to 1 (highest vulnerability) across U.S. census tracts. Higher overall SVI scores indicate increased social vulnerability, with elevated scores for individual themes reflecting greater vulnerability specific to that theme. For this analysis, raw scores were used instead of percentile rankings to better capture the contributions of individual variables to social vulnerability.

**Table 1.** Summary of the SVI themes and the attribute variables (Centers for Disease Control and Prevention/ Agency for Toxic Substances and Disease Registry/ Geospatial Research, 2020).

| **SVI Theme** | **Variable** | **Description** |
| --- | --- | --- |
| Socioeconomic Status (SES) | Below Poverty | Percentage of persons below the federal poverty level. |
|  | Unemployment | Percentage of civilian population aged 16+ who are unemployed. |
| | Housing Cost Burden | Percentage of households where housing costs exceed 30% of income for those earning < \$35,000 annually. |
|  | No High School Diploma | Percentage of persons aged 25+ without a high school diploma. |
|  | No Health Insurance | Percentage of civilian population without health insurance. |
| Household Characteristics | Aged 65 and Older | Percentage of persons aged 65 and older. |
|  | Aged 17 and Younger | Percentage of persons aged 17 and younger. |
|  | Civilian with Disability | Percentage of noninstitutionalized civilians with a disability. |
|  | Single-Parent Households | Percentage of single-parent households with children aged 18 or younger. |
|  | Limited English Proficiency | Percentage of persons aged 5+ who speak English "less than well." |
| Racial & Ethnic Minority Status | Minority Population | Percentage of persons who identify as Hispanic or Latino, Non-Hispanic Black, Asian, American Indian/Alaska Native, Native Hawaiian/Pacific Islander, multiracial, or other races. |
| Housing Type & Transportation | Multi-Unit Housing | Percentage of housing units in structures with 10 or more units. |
|  | Mobile Homes | Percentage of housing units classified as mobile homes. |
|  | Crowding | Percentage of households with more people than rooms. |
|  | No Vehicle | Percentage of households without access to a vehicle. |
|  | Group Quarters | Percentage of persons living in group quarters. |
| Overall SVI | SVI Estimate | Sum of the percentile ranks of the four SVI themes. |

### 2.3 Modeling

To model the relationship between population-weighted noise exposure, social vulnerability, and urbanization levels while accounting for spatial dependence, we fitted two spatial regression models: the spatial lag model (SLM) and the spatial error model (SEM). Moran’s I test indicated significant positive spatial autocorrelation in the residuals (I = 0.37, p < 0.001), suggesting that a standard regression approach would be inadequate due to spatial dependencies in noise exposure. Lagrange Multiplier (LM) tests identified SEM as the most appropriate model, as it provided a better fit for capturing spatial dependence. The SEM is specified as follows:

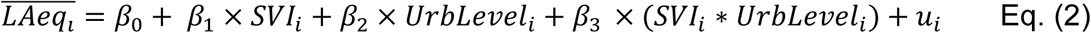

where 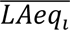 represents the population-weighted noise exposure level of census tract *i* (dBA LAeq); *SVI_i_* is a continuous variable of the overall social vulnerability index of census tract *i*; *UrbLevel_i_* is a three-level categorical variable denoting the urbanicity level of census tract *i* (0 = non-Urban Area (non-UA), 1 = Urbanized Area (UA), 2 = Highly Urbanized Area (HUA)); *SVI_i_* ∗ *UrbLevel_i_* is an interaction term between *SVI_i_* and *UrbLevel_i_*; *u_i_* is a spatially autocorrelated error term; and β_0_ - β_3_ are the regression coefficients.

As a sensitivity analysis, Census Division was introduced as an additional covariate to account for regional variations in noise exposure patterns. The adjusted SEM is specified as follows:

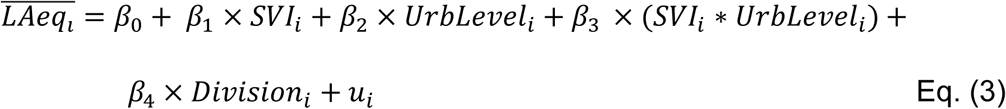

where *Division_i_* is a categorical variable representing the U.S. Census Division in which census tract *i* is located. The U.S. Census Bureau defines nine Census Divisions, which are grouped into four Census Regions: Northeast (New England and Middle Atlantic Divisions), Midwest (East North Central and West North Central Divisions), South (South Atlantic, East South Central, and West South Central Divisions), and West (Mountain and Pacific Divisions) (U.S. Census Bureau, 2021b).

### 2.4 Principal component analysis (PCA) and principal component regression (PCR)

We performed principal component analysis (PCA) to examine the underlying structure of the overall social vulnerability index (SVI) and its four thematic components: socioeconomic status (SES), household characteristics, racial and ethnic minority status, and housing type and transportation (**Table 1**). The objective was to identify the dominant patterns among these themes and determine which components explain the most variation in social vulnerability. Prior to conducting PCA, all input variables were scaled and centered to ensure comparability. Principal components were extracted from the covariance matrix of the four SVI themes, and the number of retained components was determined based on their eigenvalues and the proportion of variance explained.

The selected principal components were then used as independent variables in a principal component regression (PCR) model, estimated using a spatial error model (SEM) to account for spatial dependence. The PCR + SEM model followed the same structure as the primary SEM model (Eq. (2)), with principal components replacing the overall SVI variable. A three-level categorical variable (*UrbLevel_i_*) was included to represent urbanization level, along with interaction terms between the principal components and urbanization level to examine whether the relationship between social vulnerability and noise exposure varied across geographic classifications. As a sensitivity analysis, Census Division also was introduced as an additional covariate to account for regional variations in noise exposure and potential interactions with the principal components, following the structure of Eq. (3).

Marginal effects within each urbanization category were calculated as linear combination of main and interaction terms. Standard errors and 95% confidence intervals were estimated using the delta method, assuming independent coefficients. For all statistical analyses, p < 0.05 indicated statistical significance. The multivariate modeling and PCA was performed using the “lm” and “stats” packages with the “prcomp” function, respectively. The spatial regression was performed with “spdep” and “spatialreg” packages. All the analyses were performed in R version 4.2.2.

## 3. Results

### 3.1 Descriptive statistics

Our analysis included 84,122 census tracts, covering 99.7% of all census tracts in the U.S., with population-weighted noise exposure level entries across the continental U.S. and the states of Alaska and Hawaii. **Table 2** summarizes the overall and theme-specific SVI values across different geographical levels. Nationwide, the overall SVIs ranged from 0 to 14.32, while for Highly Urbanized Areas (HUA), the range was narrower (1.18 to 13.81). The mean overall SVI was generally higher for HUA compared to all Urban Areas (UA), except for the household characteristics theme, which includes household composition, age, and disability-related characteristics (**Table 1**).

**Table 3** presents the median, interquartile range (IQR), and overall distribution of population-weighted noise exposure levels, stratified by overall SVI quartiles and geographical levels (Nationwide, Non-Urbanized Areas (Non-UA), Urbanized Areas (UA), and Highly Urbanized Areas (HUA)). Across all census tracts nationwide, the median population-weighted noise exposure level was 53.41 dBA, with an IQR of 52.12 – 54.99 dBA. Within UA, the median noise exposure was slightly higher at 53.43 dBA (IQR: 52.10 – 55.00 dBA), while in HUA, the median was 53.47 dBA (IQR: 51.95 – 55.39). The violin plot shows that the distribution of population-weighted noise exposure varies across geographic levels, with Highly Urbanized Areas (HUA) exhibiting the greatest spread (**Figure 1**). While most HUA tracts are characterized by relatively high noise exposure, the distribution also extends toward lower levels, reflecting greater variability within highly urbanized settings. Urbanized Areas (UA) show a more compact distribution, with fewer extreme values. In contrast, the distribution for Non-Urbanized Areas (non-UA) displays a more pronounced upper tail, suggesting that although most rural census tracts experience lower noise levels, some are subject to elevated exposures. These higher exposures likely correspond to tracts located near major transportation corridors traversing otherwise low-density areas.

**Table 2.** Summary of the theme-specific and the overall SVI of different geographical area.

| Geographical level | Min | Median<br>(IQR) | Mean<br>(SD) | Max |
| --- | --- | --- | --- | --- |
| **SES theme** |  |  |  |  |
| Nationwide | 0 | 2.48<br>(1.70) | 2.49<br>(1.08) | 4.96 |
| Non Urban Area | 0 | 2.56<br>(1.26) | 2.54<br>(0.87) | 4.95 |
| All Urbanized Areas | 0 | 2.42<br>(1.79) | 2.45<br>(1.11) | 4.96 |
| Highly Urbanized Area | 0 | 2.47<br>(2.01) | 2.49<br>(1.18) | 4.95 |
| **Household characteristics theme** |  |  |  |  |
| Nationwide | 0 | 2.48<br>(0.85) | 2.46<br>(0.65) | 4.78 |
| Non Urban Area | 0 | 2.53<br>(0.76) | 2.53<br>(0.58) | 4.50 |
| All Urbanized Areas | 0 | 2.46<br>(0.90) | 2.45<br>(0.68) | 4.78 |
| Highly Urbanized Area | 0 | 2.45<br>(0.87) | 2.42<br>(0.66) | 4.52 |
| **Racial & ethnic minority status theme** |  |  |  |  |
| Nationwide | 0 | 0.50<br>(0.50) | 0.50<br>(0.29) | 1.00 |
| Non Urban Area | 0 | 0.25<br>(0.42) | 0.33<br>(0.27) | 1.00 |
| All Urbanized Areas | 0 | 0.48<br>(0.44) | 0.49<br>(0.27) | 1.00 |
| Highly Urbanized Area | 0 | 0.68<br>(0.41) | 0.64<br>(0.25) | 1.00 |
| **Housing type & transportation theme** |  |  |  |  |
| Nationwide | 0 | 2.27<br>(1.22) | 2.22<br>(0.87) | 4.70 |
| Non Urban Area | 0 | 2.39<br>(1.08) | 2.37<br>(0.76) | 4.52 |
| All Urbanized Areas | 0 | 2.16<br>(1.24) | 2.13<br>(0.87) | 4.70 |
| Highly Urbanized Area | 0 | 2.30<br>(1.31) | 2.22<br>(0.92) | 4.63 |
| **Overall SVI** |  |  |  |  |
| Nationwide | 0 | 7.62<br>(3.36) | 7.67<br>(2.25) | 14.32 |
| Non Urban Area | 0 | 7.74<br>(2.57) | 7.76<br>(1.87) | 14.10 |
| All Urbanized Areas | 1.06 | 7.42<br>(3.51) | 7.53<br>(2.32) | 14.32 |
| Highly Urbanized Area | 1.18 | 7.78<br>(3.87) | 7.78<br>(2.42) | 13.81 |

**Table 3:** Summary of the population-weighted transportation noise exposure level for Nationwide, Urbanized Areas, and Highly Urbanized Area.

| Overall SVI<br>quartile<br>(cut point) | Population-weighted noise exposure level (dba LAeq) |  |  |  |
| --- | --- | --- | --- | --- |
|  | Median<br>(25% and 75% percentile) |  |  |  |
|  | Nationwide<br>(N* = 83,437) | Non-UA<br>(N = 21,434) | Urbanized<br>Areas<br>(N = 33,054) | Highly<br>Urbanized Area<br>(N = 28,949) |
| Q1 (6.00) | 53.49<br>(52.29, 55.01) | 53.49<br>(52.53, 54.83) | 53.56<br>(52.34, 55.06) | 53.38<br>(52.08, 55.08) |
| Q2 (7.62) | 53.48<br>(52.26, 54.98) | 53.39<br>(52.40, 54.58) | 53.53<br>(52.25, 55.07) | 53.54<br>(52.09, 55.39) |
| Q3 (9.37) | 53.41<br>(52.12, 54.95) | 53.34<br>(52.31, 54.49) | 53.45<br>(52.08, 55.02) | 53.51<br>(51.90, 55.50) |
| Q4 (14.32) | 53.24<br>(51.80, 55.00) | 53.10<br>(51.97, 54.35) | 53.10<br>(51.70, 54.78) | 53.51<br>(51.72, 55.57) |
| All | 53.41<br>(52.12, 54.99) | 53.34<br>(52.30, 54.55) | 53.43<br>(52.10, 55.00) | 53.47<br>(51.95, 55.39) |

\*Total number of the census tract included in the analysis.

**Figure 1.**
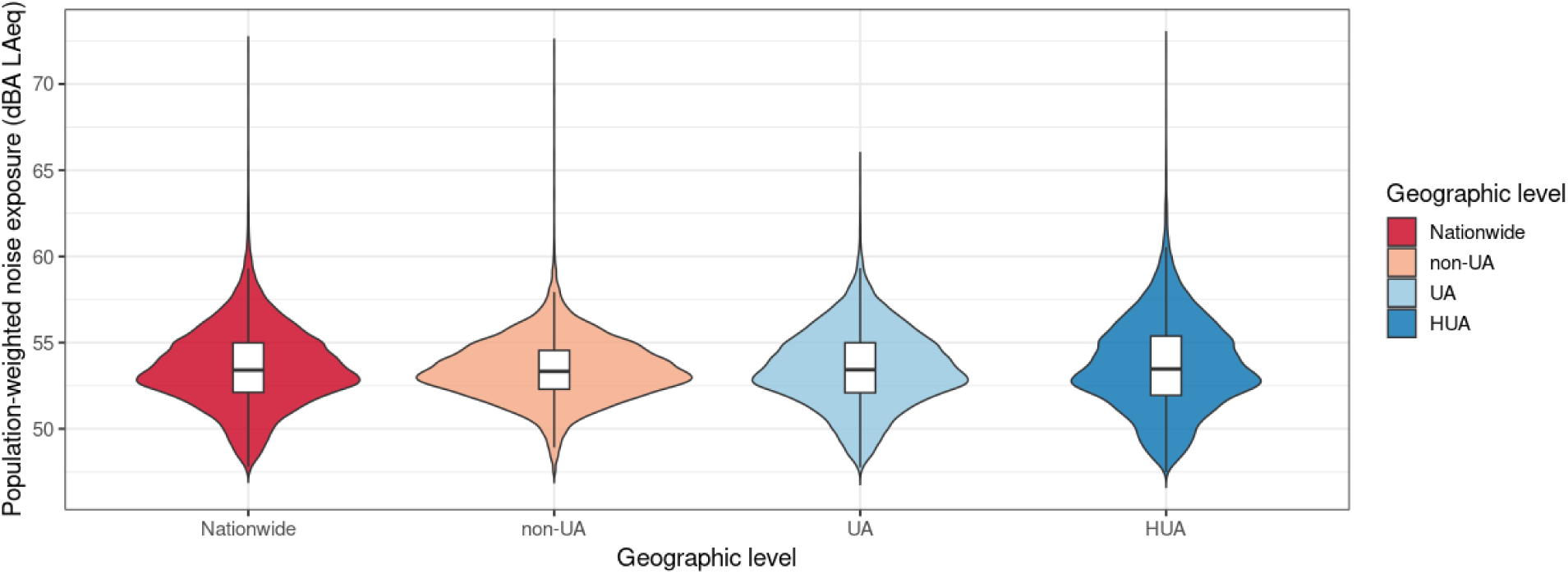
Distribution of population-weighted noise exposure across geographic levels.

Figures 2 **and 3** illustrate spatial distribution of the ranked SVI and population-weighted transportation noise exposure level in the continental U.S., respectively (maps of the states of Alaska and Hawaii for ranked SVI and population-weighted transportation noise exposure level are presented in **Figures S1 and S2, respectively**). Generally, we observe pockets of elevated SVI in all states. There are notable parts of the country though, we find regions of census tracts with higher SVI rankings, such as in the states of Alabama, Kentucky, Mississippi, and Tennessee (Centers for Disease Control and Prevention, 2023). In comparison, elevated population-weighted transportation noise exposure levels are observed along the networks of both inter and intrastate rail and roadway lines, and tend to be clustered in major urban areas, which is particularly noticeable for cities in the Midwestern and Eastern U.S.

**Figure 2.**
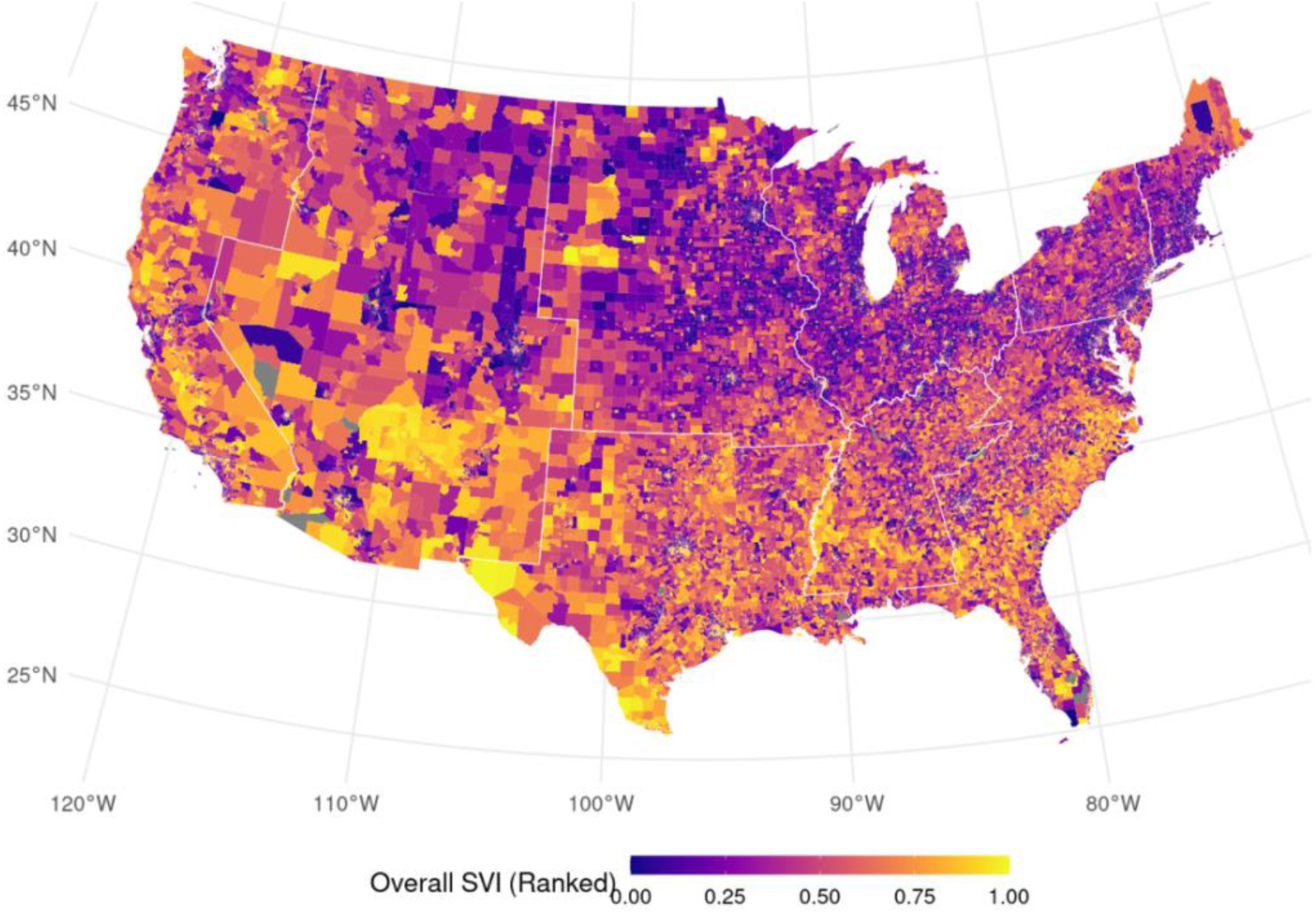
Map of the ranked Social Vulnerability Index (SVI) in the Continental U.S. with Census Division boundaries. Maps for the states of Alaska and Hawaii are shown in Figure S1.

**Figure 3.**
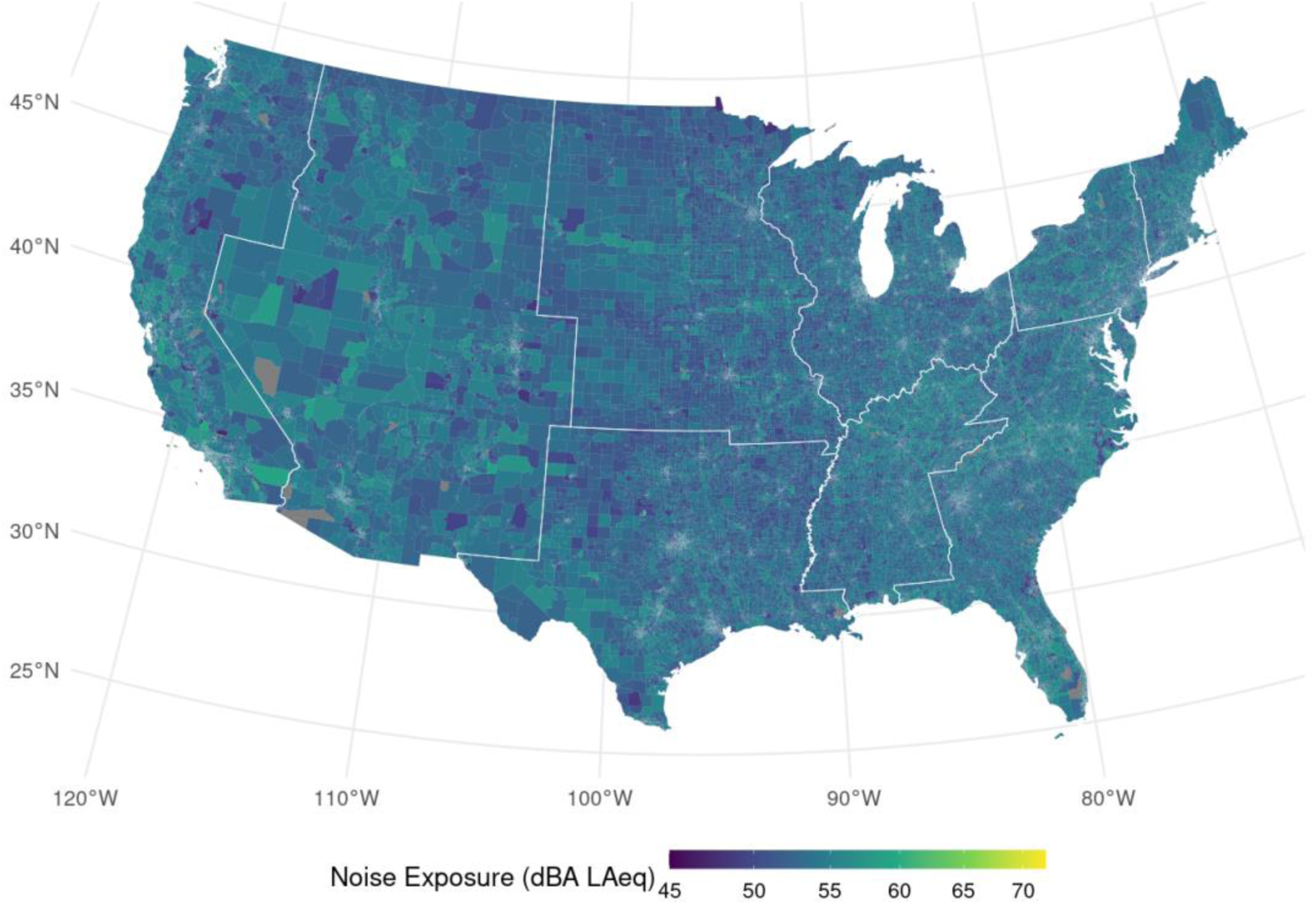
Population-weighted transportation noise exposure (dBA LAeq) across continental U.S. with Census Division boundaries. Maps for the states of Alaska and Hawaii are shown in Figure S2.

### 3.2 Associations between SVI and Noise Exposure

The SEM results (**Table 4**) indicate significant spatial dependence in noise exposure across census tracts, as reflected by the estimated spatial error coefficient (λ = 0.661, 95% CI: 0.53 to 0.80, p < 0.001). This suggests that unobserved spatial processes contribute to transportation noise patterns. The SEM results further reveal that the association between social vulnerability (SVI) and population-weighted transportation noise exposure varies by urbanization level.

The marginal effect of SVI was negative in non-urban areas (β = -0.060, 95% CI: -0.076 to -0.044, p < 0.001), indicating that census tracts in less urbanized settings with higher SVI tended to have lower noise exposure levels. In urbanized areas (UA), the marginal effect was positive but weaker (β = 0.022, 95% CI: -0.003 to 0.047), suggesting a slight increase in noise exposure with higher SVI. In highly urbanized areas (HUA), the marginal effect was stronger (β = 0.061, 95% CI: 0.034 to 0.088), indicating that census tracts in these areas with higher SVI were associated with significantly higher noise exposure.

Urbanization level itself was also significantly associated with noise exposure, with both UA and HUA having lower intercept values compared to non-UA (β = -0.595, p < 0.001 for UA; β = -0.760, p < 0.001 for HUA). These findings suggest that urbanization moderates the relationship between social vulnerability and noise exposure, with opposing trends observed in rural versus urban settings. A sensitivity analysis adjusting for Census Division (**Table S1**) confirmed these findings, showing minimal regional variation. The results indicate that higher social vulnerability is linked to increased noise exposure in urbanized regions, while in rural areas, higher social vulnerability is associated with lower exposure.

**Table 4.** Summary of the coefficients of the SEM model based on Eq. (2).

| Variable | Coef | SE | 95%CI | p-value |
| --- | --- | --- | --- | --- |
| Intercept | 53.803 | 0.069 | 53.668 to 53.939 | <0.001 |
| Overall SVI | -0.060 | 0.008 | -0.076 to -0.044 | <0.001 |
| UL<sup>a</sup> |  |  |  |  |
| UA | -0.595 | 0.081 | -0.754 to -0.437 | <0.001 |
| HUA | -0.760 | 0.090 | -0.936 to -0.584 | <0.001 |
| Overall SVI: UL |  |  |  |  |
| **Overall SVI: UA** | 0.082 | 0.010 | 0.062 to 0.101 | <0.001 |
| **Overall SVI: HUA** | 0.121 | 0.011 | 0.100 to 0.141 | <0.001 |

<sup>a</sup> The group non-urbanized area (non-UA) is used as the reference group.
Definition of abbreviations: Coef: coefficient; SE: standard error; 95% CI: 95% confidence interval; SVI: social vulnerability index; UA: urbanized area; HUA: highly urbanized area.

### 3.3 Associations between SVI Components and Noise

The PCA yielded two principal components (PCs) that together accounted for 75.2% of the variance in the SVI themes across census tracts in the U.S. (Scree plot shown in **Figure S3**). Figure 4 illustrates the principal component loadings for each SVI theme. The first component (PC1) exhibits strong positive loadings across all four SVI themes, indicating that it captures broad social vulnerability across multiple dimensions, including socioeconomic status, household characteristics, racial/ethnic minority status, and housing type/transportation. This aligns with the overall SVI measure.

The second component, PC2, exhibits a strong positive loading for racial/ethnic minority status, a moderate loading for household characteristics, and negative loadings for both housing type/transportation and socioeconomic status. This suggests that census tracts with high PC2 scores tend to have a large proportion of racial/ethnic minority populations and some household vulnerabilities, such as a higher proportion of children, older adults, or individuals with disabilities. However, these areas are less likely to experience severe socioeconomic disadvantage, such as poverty, unemployment, or limited access to education and resources. They are also less likely to exhibit housing/transportation-related vulnerabilities, such as high-density multi-unit housing, crowding, or lack of vehicle access, compared to areas with high PC1 scores (**Table 1**).

**Figure 4.**
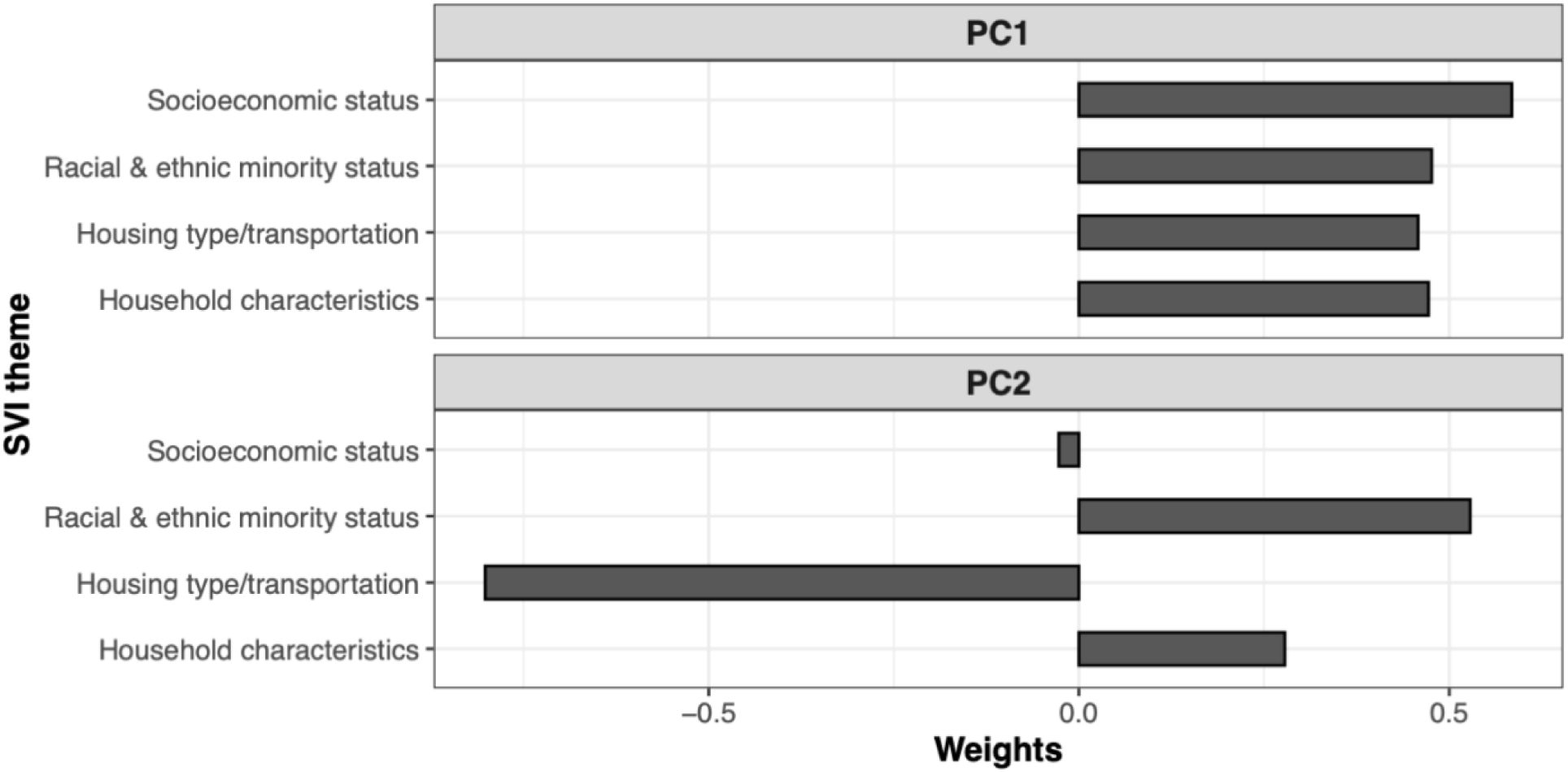
Principal component factor loadings for the first two features, which explain 75.2% of the variation in SVI themes.

The SEM+PCR results (**Table 5**) revealed distinct patterns in the associations between principal components of social vulnerability and transportation noise exposure. Census tracts with higher overall social vulnerability (PC1) had significantly lower noise exposure in non-urbanized areas (marginal effect: β = -0.097, 95% CI: -0.122 to -0.072). However, in urbanized and highly urbanized areas, this relationship reversed, with PC1 being associated with higher noise exposure (marginal effect in UA: β = 0.008, 95% CI: -0.032 to 0.048; marginal effect in HUA: β = 0.068, 95% CI: 0.027 to 0.109). The positive and statistically significant interaction terms between PC1 and urbanization (PC1 × UA, β = 0.105, p < 0.001; PC1 × HUA, β = 0.165, p < 0.001) confirm that the association between social vulnerability and noise exposure changes across levels of urbanization, shifting from negative from negative rural areas to positive in urban centers.

For PC2, the main effect was not significant (β = -0.019, p = 0.334), indicating no clear association between PC2 and noise exposure overall. However, the marginal effects of PC2 suggest a different trend in urban settings, where census tracts with high PC2 scores—characterized by a large proportion of racial/ethnic minority populations but lower socioeconomic and housing/transportation vulnerability—tend to experience lower noise exposure (marginal effect in UA: β = -0.133, 95% CI: -0.196 to -0.70; marginal effect in HUA: β = -0.135, 95% CI: -0.199 to -0.071). The negative and significant interaction terms (PC2 × UA, β = -0.114, p < 0.001; PC2 × HUA, β = -0.116, p < 0.001) suggest that while these communities may be socially vulnerable in some respects—such as having higher proportion of racial/ethnic minority populations—they may also have protective factors (e.g., better housing conditions or greater vehicle access) that mitigate noise exposure risks in urban settings.

A sensitivity analysis adjusting for Census Division (**Table S2**) further confirmed the role of urbanization in modifying the relationship between different vulnerability factors and noise exposure. For PC2 tracts where vulnerability is primarily driven by racial/ethnic minority status and household characteristics, the relationship between SVI and noise exposure became more negative with increasing urbanization, suggesting that these communities experience lower transportation noise exposure in more urbanized settings. While PC2 effects were not statistically significant in the main model, they became significant in the sensitivity analysis, suggesting that regional factors play a key role in shaping noise disparities for these communities. This demonstrates that SVI-driven noise disparities for PC2 tracts vary by Census Division and the importance of regional context in understanding environmental noise exposure patterns beyond what is captured by the overall SVI alone.

**Table 5.** Summary of the coefficients of the SEM+PCR model based on eq. (2). (lambda = 0.656, p < 0.001)

| Variable | Coef | SE | 95%CI | p-value |
| --- | --- | --- | --- | --- |
| **Intercept** | 53.336 | 0.032 | 53.272 to 53.399 | <0.001 |
| **PC1** | -0.097 | 0.013 | -0.122 to -0.072 | <0.001 |
| **PC2** | -0.019 | 0.020 | -0.059 to 0.020 | 0.3373 |
| **UL<sup>a</sup>** |  |  |  |  |
| **UA** | 0.043 | 0.037 | -0.028 to 0.115 | 0.2351 |
| **HUA** | 0.197 | 0.043 | 0.112 to 0.282 | <0.001 |
| **PC1:UL** |  |  |  |  |
| **PC1: UA** | 0.105 | 0.016 | 0.074 to 0.136 | <0.001 |
| **PC1: HUA** | 0.165 | 0.017 | 0.132 to 0.198 | <0.001 |
| **PC2:UL** |  |  |  |  |
| **PC2:UA** | -0.114 | 0.025 | -0.163 to -0.065 | <0.001 |
| **PC2:HUA** | -0.116 | 0.026 | -0.168 to -0.064 | <0.001 |

<sup>a</sup> The group non-urbanized area (non-UA) is used as the reference group.
Definition of abbreviations: Coef: coefficient; SE: standard error; 95% CI: 95% confidence interval; SVI: social vulnerability index; UA: urbanized area; HUA: highly urbanized area

## 4. Discussion

While previous studies have assessed race/ethnicity and SES indicators as independent risk factors for noise exposure, this is the first study that has investigated the relationship between the SVI and transportation noise exposure level across the U.S. Our approach is novel and different from previous studies of transportation noise inequality in our consideration of the SVI — a composite vulnerability index, consideration of principal components that underlie the variation in social vulnerability, and consideration of the possible interactive effects of social vulnerability in highly urban vs. non-urban areas. Our results revealed there is a significant interaction between census tract-level overall social vulnerability and high urbanization levels in influencing transportation noise exposure. We discovered that the urbanization level of census tracts in the U.S. is a key factor on impacting population’s transportation noise exposure. For census tracts that are within the highly urbanized area (i.e., the top 5% Urban Areas as defined by U.S. Census Bureau), having higher overall SVI is associated with higher transportation noise exposure. In contrast, for census tracts that are in less urbanized areas, higher overall SVI is associated with lower transportation noise exposure. We discuss the implications for environmental justice, and for interventions aimed at mitigating the adverse effects of transportation noise on vulnerable communities.

Decomposing the overall SVI by PCA, our findings reveal the role of different sociodemographic factors in shaping overall social vulnerability across the U.S., as well as the relationship between different aspects of social vulnerability and transportation noise exposure. Although the overall SVI framework is already structured to encompass four main themes (socioeconomic status, household characteristics, racial and ethnicity minority status, and housing/transportation factors), the scoring method of adding the four sub-scores, does not account for correlations that may exist within the multidimensional space. Our use of PCA considers these correlations and reduces the dimensionality of these four themes into two key principal components that explain most of the underlying variation in the SVI themes. The extracted principal component features have semantic meaning: while both features represent neighborhoods with high proportions of racial/ethnic minorities and vulnerable household characteristics, one is more vulnerable than the other with respect to SES and housing/transportation factors.

PC1, which represents census tracts with low scores on all four SVI themes, essentially captured the area with high overall vulnerability as the original overall SVI. Echoing our multivariate regression analysis and previous studies (Casey et al., 2017, Simon et al., 2022), the PCR results highlight that census tracts within highly urbanized area with elevated PC1 scores, namely census tracts with population of minor race ethnicity and with higher social vulnerability in terms of housing, household characteristics, and transportation, are associated with higher transportation noise exposure level, whereas census tracts that are within the less urbanized area, higher overall SVI are associated with lower transportation noise exposure level (Figure 5(a)).

The extracted PC2 represents census tracts that still have high proportions of racial/ethnic minority populations and household vulnerability, but are relatively less vulnerable with respect to SES and housing type/transportation themes. PCR outcomes suggest that the level of urbanization has an opposite interactive effect for those living in these census tracts; in less urbanized areas racial/ethnic minority communities tend to be more exposed to transportation noise. Conversely, for these specific census tracts located in highly urbanized areas, racial/ethnic minority communities who are less vulnerable with respect to SES and housing/transportation-related factors tend to be less exposed to transportation noise (Figure 5(b)). This highlights the influential role of SES and accessibility of individual housing and vehicles in shaping the vulnerability of populations to transportation noise, as high SES and individual housing and vehicles may serve as a protective factor against noise exposure in urbanized settings for race and ethnic minorities. The underlying factors contributing to SES themes encompass education level, income, employment status, housing costs, and healthcare insurance ownership (**Table 1**). Existing research has demonstrated the complex relationship between race/ethnicity and SES, with variations in SES distribution evident among different racial and ethnic groups (Williams et al., 2010). Notably, in the US, previous studies have found that household income has the potential to alter risks that different racial/ethnic minority populations face from environmental hazards (Morello-Frosch et al., 2001).

In line with previous studies, our findings emphasize the pivotal role of urbanization in shaping transportation noise. Understanding the differences in noise exposures between vulnerable populations living in highly urban areas versus those that do not live in these areas is important in the United States, where recent demographic data indicate increasing proportions of racial/ethnic minorities not only in rural counties, but also in urban counties (Parker et al., 2018). Urban areas tend to have heightened density and prevalence of transportation infrastructure, including airports, railways, and roads, contribute significantly to increased noise levels (Seto et al., 2007). Factors such as traffic congestion and proximity to transportation hubs further exacerbate noise exposure in these settings (Vijay et al., 2015). Furthermore, land-use patterns in urbanized area, characterized by a mix of residential, commercial, and industrial activities, could also amplify the impact of transportation noise exposure (Yildirim and Arefi, 2021, King et al., 2012, Han et al., 2017). The clustering of diverse human activities and population within urban settings leads to cumulative impacts on transportation noise exposure. As such, from a public health perspective, understanding the role of urbanization in mediating the relationship between social vulnerability and noise exposure is imperative in developing urban planning strategies aimed at mitigating the adverse effects of transportation noise, particularly on vulnerable populations residing in highly urbanized area. The results of this study compliment prior studies examining different outcomes with the SVI, which concludes communities with high overall SVI are more vulnerable to natural disasters and environmental hazards (Ramesh et al., 2022, Lehnert et al., 2020, Lieberman-Cribbin et al., 2020, Dasgupta et al., 2020), while adding granularity by identifying the distinct impacts of individual SVI components on transportation noise exposure.

Our findings complement previous studies that have considered race/ethnicity and SES as independent risk factors for noise exposure. Notably, a U.S. wide study of modeled L_50_ noise levels observed higher noise exposures for census block groups with higher proportions of nonwhite and lower SES populations (Casey et al., 2017). Racial segregation quantified via the neighborhood dissimilarity index was also found to be associated with noise exposure. Furthermore, that study also observed multiple SES indicators, such as poverty, unemployment, linguistic isolation, and educational attainment were associated with noise exposure. Similar findings were also reported in a nationwide study of transportation-related noise exposures, which found that roadway and aviation noise tended to be higher in census tracts with greater proportions of nonwhite populations, as well as neighborhood deprivation (Collins et al., 2020). This study adjusted for urbanicity (the Rural-Urban Commuting Area (RUCA) codes) in multivariate models, and found as we would expect, higher noise exposures in urban areas, but did not consider the potential interaction between social vulnerability and urbanicity on noise exposures. Similar findings of environmental inequality in the associations between race/ethnicity, SES, and noise exposure have also been reported for North American studies conducted in specific regions, including for modeled roadway noise exposures for public schools in Texas (Chakraborty and Aun, 2023), modeled road traffic noise in Montreal (Carrier et al., 2016) modeled roadway and aviation related noise in Georgia (Cohen et al., 2019) and the Twin Cities of Minnesota (Nega et al., 2013), and 5-minute measured daytime noise levels in Chicago (Huang et al., 2021). Furthermore, neither of these previous national studies were able to assess differences in exposures by SES level for racial/ethnic minority groups.

In contrast to the previous studies, mixed findings have been reported for social vulnerability and noise in non-North American studies. In studies conducted in the Netherlands, lower income levels were found to be associated with noise exposures in both the Rijnmond and Schiphol Airport areas (Kruize et al., 2007b, Kruize et al., 2007a). Similarly, in Hong Kong, exposure to road noise was found to be associated with socioeconomic disadvantage (Lam and Chan, 2008). Also, in the UK, Black and low-income populations were found to live in Birmingham neighborhoods with higher roadway noise (Brainard et al., 2004). However, in London, roadway traffic noise was found to be low amongst those with high income and amongst Asian populations, but aviation noise was found to be experienced by high income and White populations. A study of noise in Paris, also observed an opposite relationship between noise and SES indicators: higher noise was found in neighborhoods with higher property values and higher educational attainment (Havard et al., 2011). These mixed findings reinforce the need for continued research on the social inequalities of environmental noise exposures, particularly with respect to region-specific contexts (Dreger et al., 2019).

Our analysis has shared some limitations with previous studies of noise exposure inequality. First, the primary outcome in our analysis is based on the LAeq noise metric, which is the 24-hour average noise level, without incorporating any penalties or weighting based on the time of day. Transportation noise can exhibit variations throughout the day due to traffic patterns, potentially causing more disturbance during nighttime hours. A recent study highlighted that LAeq noise metric consistently underestimates noise levels compare to penalized noise metrics by approximately 3 dBA (Seto and Huang, 2023a). Therefore, for assessing health impacts, it is essential to consider penalized noise metrics such as Lden and Ldn, which account for time-specific weighting and prevent estimation bias. This limitation arises from the nature of the transportation noise dataset employed in our study, where the modeled transportation noise level was based on LAeq. Also, while the LAeq is a common environmental noise metric, it is difficult to compare our findings with those of a previous national study, which used the less common L_50_ metric (Casey et al., 2017). Secondly, our study utilized transportation data from 2020 obtained from the BTS and a prior study, along with 2020 CDC/ATSDR SVI data. Statistics have shown decreased traffic noise levels during the 2020 COVID-19 pandemic compared to pre-pandemic periods in the US (U.S. Department of Transporation, 2023). Future research aimed at investigating trends in transportation noise data and SVI before, during, and after the major events that alter traffic patterns could provide valuable insights into the dynamics of the relationship between transportation noise exposure and social vulnerability. Third, the presence of moderate positive spatial autocorrelation in noise exposures at the census tract level suggests that unmeasured spatial processes may influence the observed noise levels. Like other studies that have considered noise exposure equity, we employed spatial regression models to account for this autocorrelation in the relationship between overall SVI and noise exposure. Finally, our study did not explicitly focus on children as a vulnerable population group, a limitation shared with many prior studies. While previous research, especially those conducted in school settings, has emphasized the need to address children’s exposure to transportation noise, less work has been done to examine children’s exposures outside of school (Collins et al., 2019). Children may experience distinct exposure patterns due to their mobility and time spent at home or in transit, which can differ significantly from adult populations. Future research should address this gap by explicitly investigating children’s noise exposures across various environments, particularly outside of school (Chakraborty and Aun, 2023), to better understand the cumulative impact of transportation noise on this vulnerable population group.

Despite these limitations, our study has notable strengths. To our knowledge, it is the first to explore the structure of SVI and its connection transportation noise exposure. We applied PCA to understand how overall SVI depends on different sociodemographic and socioeconomic factors, providing a detailed perspective on the SVI’s underlying structure. Our use of PCR further leverages the PCA results by integrating the identified principal components as independent variables in a regression model. The results of our study reveal how specific social vulnerability components, inherently tied to environmental justice considerations, relate to transportation noise exposure.

Additionally, our exploration on the interaction effects across different urbanization levels of the census tracts allow us to identify whether the impact of social vulnerability on noise exposure varies depending on the level of urbanization, emphasizing the need for addressing environmental justice concerns and informing targeted interventions to mitigate the adverse effects of transportation noise on vulnerable communities.

## 5. Conclusion

In this study, we leverage publicly available national transportation noise exposure and the SVI to explore the relationship between social vulnerability and transportation noise exposure across the U.S. Our results highlight the influence of specific social vulnerability components on transportation noise exposure across different urbanization strata. The findings of this study expand the current knowledge on the link between social vulnerability and environmental hazard exposures, highlighting the multifaceted nature of environmental justice and the need of tailored strategies to address the disparate impacts of transportation noise on public health.

## Data Availability

All data produced in the present study are available upon reasonable request to the authors

