## Supplemental Materials for "Social Inequities in Transportation Noise Exposures in the United States: Urbanicity Modifies that Relationship Between Social Vulnerability and Noise Exposure"

(a)

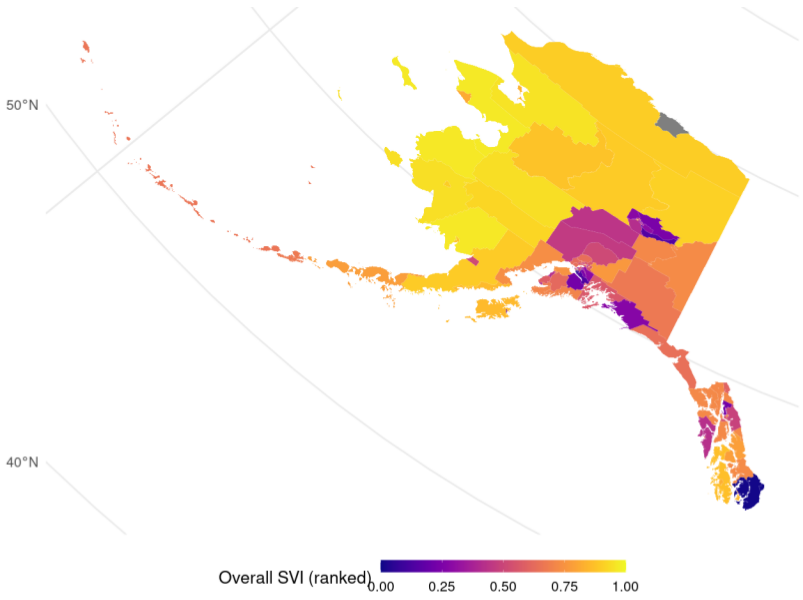


(b)

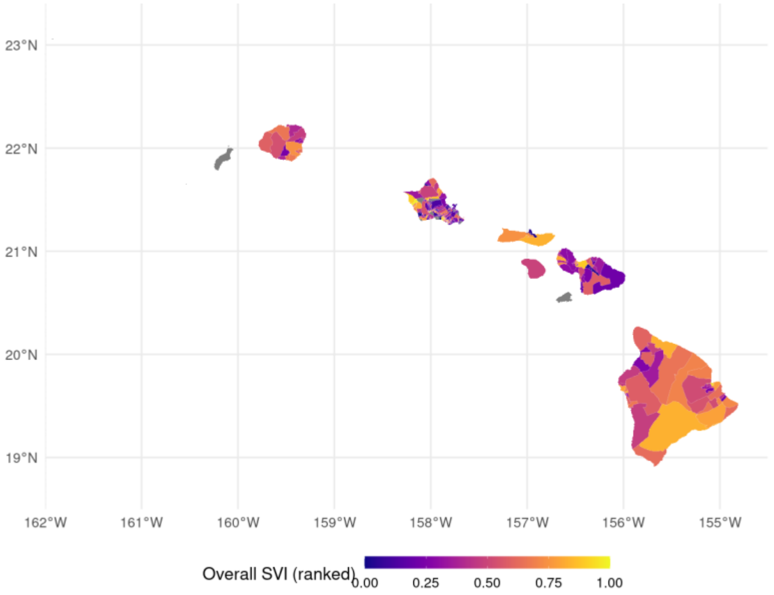


**Figure S1. Maps of the ranked Social Vulnerability Index in the states of Alaska and Hawaii. Gray areas indicate missing data.**

(a)

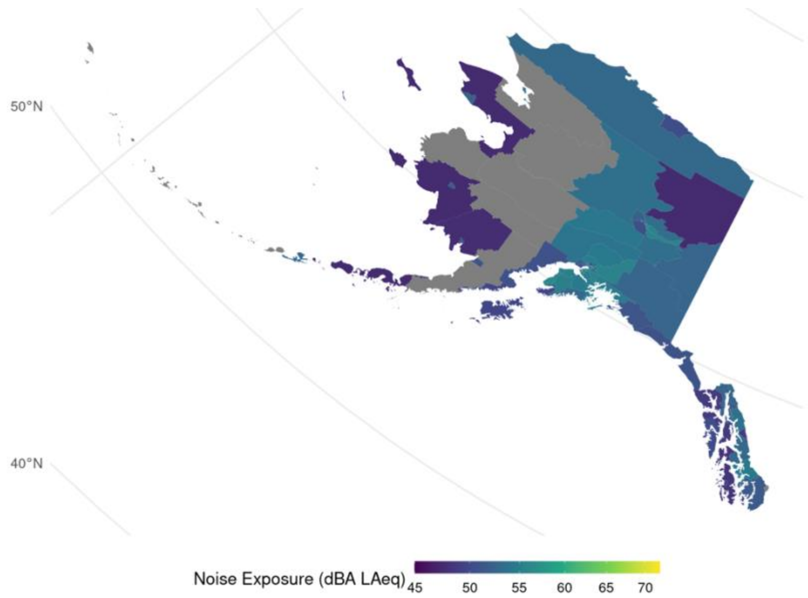


(b)

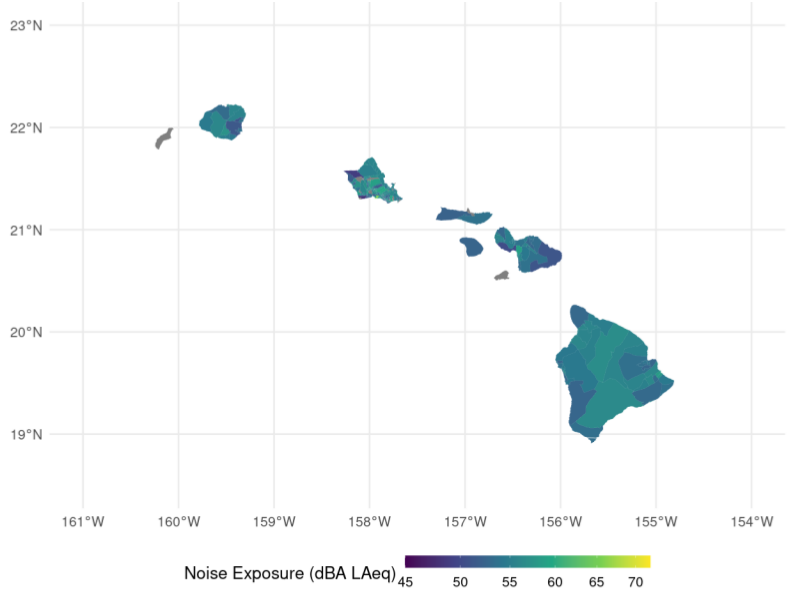


**Figure S2. Maps of the population-weighted transportation noise exposure level in the states of Alaska and Hawaii. Gray areas indicate missing data.**

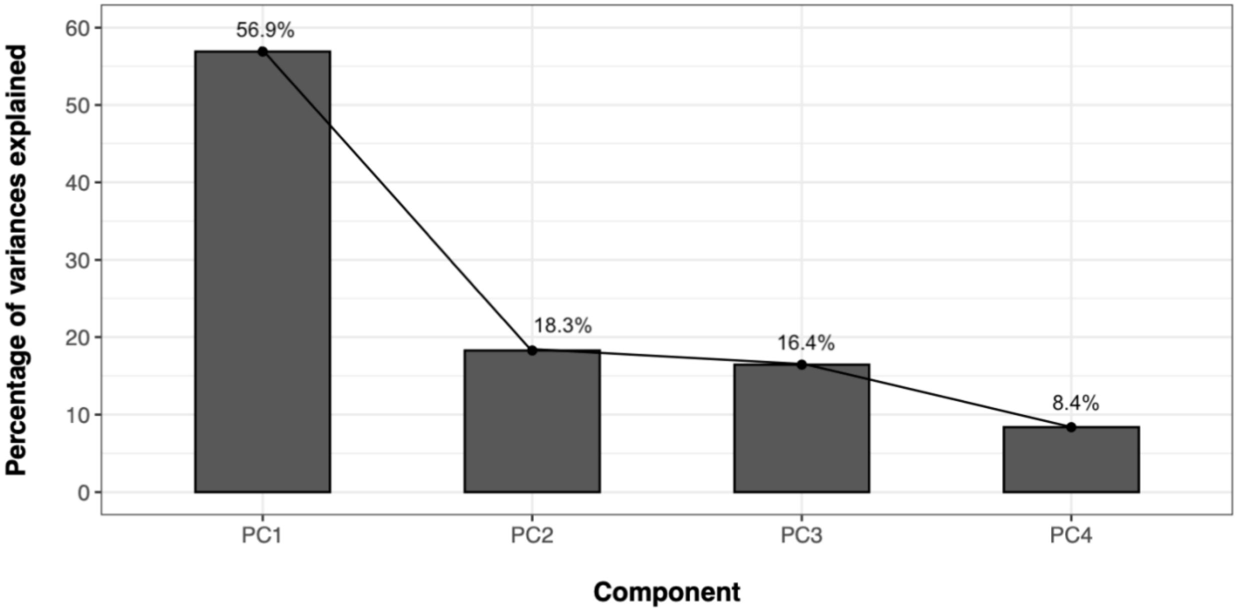


**Figure S3. Scree plot of the PCA.**

**Table S1. Summary of the coefficients of the SEM sensitivity model based on eq. (3). (lambda = 0.649, p < 0.001)**

| Variable | Coef | SE | 95%CI | p-value |
|---|---|---|---|---|
| **Intercept** | 54.159 | 0.079 | 54.005 to 54.314 | <0.001 |
| **Overall SVI** | -0.064 | 0.008 | -0.080 to -0.048 | <0.001 |
| **UL[a]** | | | | |
| **UA** | -0.619 | 0.081 | -0.777 to -0.46 | <0.001 |
| **HUA** | -0.772 | 0.090 | -0.948 to -0.597 | <0.001 |
| **Division** | | | | |
| **ENC** | -0.483 | 0.062 | -0.603 to -0.362 | <0.001 |
| **ESC** | -0.079 | 0.081 | -0.238 to 0.080 | 0.331 |
| **MA** | -0.804 | 0.065 | -0.932 to -0.677 | <0.001 |
| **MTN** | -0.224 | 0.078 | -0.377 to -0.070 | <0.05 |
| **NE** | -0.203 | 0.093 | -0.386 to -0.020 | <0.05 |
| **PAC** | 0.139 | 0.063 | 0.015 to 0.262 | <0.05 |
| **WNC** | -0.835 | 0.079 | -0.990 to -0.681 | <0.001 |
| **WSC** | -0.523 | 0.066 | -0.653 to -0.393 | <0.001 |
| **Overall SVI: UL** | | | | |
| **Overall SVI: UA** | 0.082 | 0.010 | 0.062 to 0.101 | <0.001 |
| **Overall SVI: HUA** | 0.122 | 0.011 | 0.101 to 0.142 | <0.001 |

[a] The group non-urbanized area (non-UA) is used as the reference group.
Definition of abbreviations: Coef: coefficient; SE: standard error 95% CI: 95% confidence interval; SVI: social vulnerability index; UA: urbanized area; HUA: highly urbanized area

**Table S2. Summary of the coefficients of the SEM+PCR sensitivity model based on eq. (3). (lambda = 0.648, p < 0.001)**

| Variable | | Coef | SE | 95%CI | p-value |
|---|---|---|---|---|---|
| **Intercept** | | 53.668 | 0.049 | 53.573 to 53.764 | <0.001 |
| **PC1** | | -0.111 | 0.013 | -0.137 to -0.086 | <0.001 |
| **PC2** | | -0.046 | 0.020 | -0.086 to -0.006 | <0.05 |
| **UL** [a] | | | | | |
| | **UA** | 0.029 | 0.036 | -0.041 to 0.100 | 0.417 |
| | **HUA** | 0.207 | 0.044 | 0.122 to 0.293 | <0.001 |
| **Division** | | | | | |
| | **ENC** | -0.516 | 0.062 | -0.637 to -0.395 | <0.001 |
| | **ESC** | -0.092 | 0.081 | -0.250 to 0.067 | 0.258 |
| | **MA** | -0.847 | 0.065 | -0.975 to -0.720 | <0.001 |
| | **MTN** | -0.232 | 0.078 | -0.385 to -0.079 | <0.01 |
| | **NE** | -0.264 | 0.093 | -0.447 to -0.081 | <0.01 |
| | **PAC** | 0.136 | 0.063 | 0.013 to 0.259 | <0.05 |
| | **WNC** | -0.894 | 0.079 | -1.049 to -0.739 | <0.001 |
| **PC1:UL** | | | | | |
| | **PC1: UA** | 0.109 | 0.016 | 0.078 to 0.140 | <0.001 |
| | **PC1: HUA** | 0.172 | 0.017 | 0.139 to 0.205 | <0.001 |
| **PC2:UL** | | | | | |
| | **PC2:UA** | -0.098 | 0.025 | -0.147 to -0.049 | <0.001 |
| | **PC2:HUA** | -0.098 | 0.026 | -0.150 to -0.046 | <0.001 |

[a] The group non-urbanized area (non-UA) is used as the reference group.
Definition of abbreviations: Coef: coefficient; SE: standard error 95% CI: 95% confidence interval; SVI: social vulnerability index; UA: urbanized area; HUA: highly urbanized area
